# SARS-CoV-2 seroprevalence on the north coast of Peru: A cross-sectional study after the first wave

**DOI:** 10.1101/2022.09.07.22279669

**Authors:** Luz M. Moyano, Angie K. Toledo, Jenny Chirinos, Percy Mc Quen Vilchez Barreto, Sofia Cavalcanti, Ricardo Gamboa, Jhon Ypanaque, Mauro Meza, Sheila Noriega, Victor Herrera, Edgar Bazan, Alexandra Requena, Henry Silva, Harold Burgos, Franco León-Jimenez, Group of Neuroepidemiology and Science of Life of Peru

## Abstract

**Background:** The devastating repercussions of COVID-19 were felt in developing nations like Peru. However, few studies have been conducted in these countries. To make good decisions about public health, it is important to understand how the disease is spread in our area.

**Methodology/Principal findings:** An observational, cross-sectional study was performed between November 11th and November 30th, 2020. In Puerto Pizarro, one out of every four homes was invited to participate in a systematic randomized sampling. Individuals were screened for the qualitative detection of anti-SARS-CoV-2 nucleocapsid (N) protein antibodies and anti-SARS-CoV-2 spike RBD with a rapid chromatographic immunoassay. An adult of legal age was selected, and an additional molecular test (RT-PCR) was taken to look for active COVID-19 cases.

**Conclusions/Significance:** This study shows an adjusted seroprevalence of 24.72% posterior to the first wave of COVID-19 in Tumbes. When adjusted by participant characteristics, women had higher adjusted seroprevalence compared to men (213/356 vs 143/356 [28.01 % vs 21.18 %], p=0.005). More than 20% of IgG seropositive cases belong to the age group under 16 years old. Asymptomatic individuals with recent infections were 66.3% (IgM and IgM/IgG) across all age groups. No association between positive seroprevalence and water supply, water resources, or sanitation services was found. The information is relevant to the Ministry of Health’s establishment of a regional program of COVID-19 control and strategic interventions, targeting vulnerable groups and improving vaccination campaigns.

**Author summary:** COVID-19 had devastating effects on developing countries such as Peru. It’s crucial to understand the disease’s underlying distribution in our region to create useful dynamics that engage the population in prevention measures. We performed an observational, cross-sectional study between November 11th and November 30th, 2020, in Puerto Pizarro. One out of every four houses was invited to participate, and individuals were screened for the qualitative detection of anti-SARS-CoV-2 antibodies (IgG and IgM) with a rapid test. This study shows an adjusted seroprevalence of 24.72% posterior to the first wave of COVID-19 in Tumbes. Women had a higher adjusted seroprevalence compared to men (213/356 vs 143/356 [28.01 % vs 21.18 %], p=0.005). More than 20% of IgG seropositive cases belong to the age group under 16 years old. Asymptomatic individuals with recent infections were 66.3% (IgM and IgM/IgG) across all age groups. Community participation in epidemiological surveillance strategies is crucial to establish a future follow-up cohort and evaluate the medium-term sequelae of this disease.

## Introduction

In the last days of December 2019, the first cases of an unknown pneumonia were reported in the city of Wuhan, Hubei province, China (1,2). After a few weeks of research, in January 2020, the Chinese Center for Disease Control and Prevention announced that they had found a new coronavirus named SARS-CoV-2 (3,4); The World Health Organization (WHO) declared a pandemic of coronavirus disease (COVID-19), caused by the SARS-CoV-2 virus, on March 11, 2020 (5). COVID-19 had a devastating effect on developing countries. The first case in Peru was reported in March 2020 in a 25-year-old European man (6). Regional initiatives to determine the true spread of COVID-19 were conducted in Iquitos (7), Lambayeque (8), and Lima (9), with prevalence rates ranging between 29.5/1000 and 70/1000 using data from the CDC’s NOTIWEB notification program and the Ministry of Health’s (MINSA) SISCOVID software (10). In Peru, the first wave of COVID-19 ended during the first few days of December 2020. Up to December 10, 2020, Peru had registered more than 900,000 cases of COVID-19 and more than 36,000 confirmed deaths from the disease. Tumbes presented a higher mortality rate than the national mortality rate (11).

Using a large-scale community intervention called Plan Tumpis, in which the Tumbes Regional Government (GORE) and the Regional Directorate of Health (DIRESA) found symptomatic cases through active house-to-house surveillance, we did a cross-sectional study in the marginal-urban community of Puerto Pizarro to find out the SARS-CoV-2 seroprevalence, distribution by age group, and health determinants associated with this respiratory condition in residents older than 2 years old. To be able to make decisions about public health, we need to know how the disease is spread in our area.

## Materials and methods

### Ethical considerations

The study protocol and consent forms were approved by the institutional review boards of Universidad Peruana Cayetano Heredia and the Regional Directorate of the Ministry of Health (DIRESA) in Tumbes. All participants signed the informed consent (IC) in the presence of a witness. In the case of minors, the consent was given by the minor and the parents or legal guardians.

### Area and study population

Puerto Pizarro is a marginal urban port village located 13 kilometers south of Tumbes City with a population of 4,000 people (INEI 2020 census). Puerto Pizarro is well-known as the entrance to the Tumbes Mangrove Sanctuary, with a mestizo population dedicated to fishing, trading, aquaculture, and tourism (12,13). Although the residents have access to electricity, they have limited access to water and sewage services. Puerto Pizarro has a primary-care health center that is staffed by a newly graduated general practitioner, a nurse, an obstetrician, and a nursing technician on a one-year rural service. The center is open 12 hours a day from Monday to Saturday (Fig 1).

**Fig 1.**
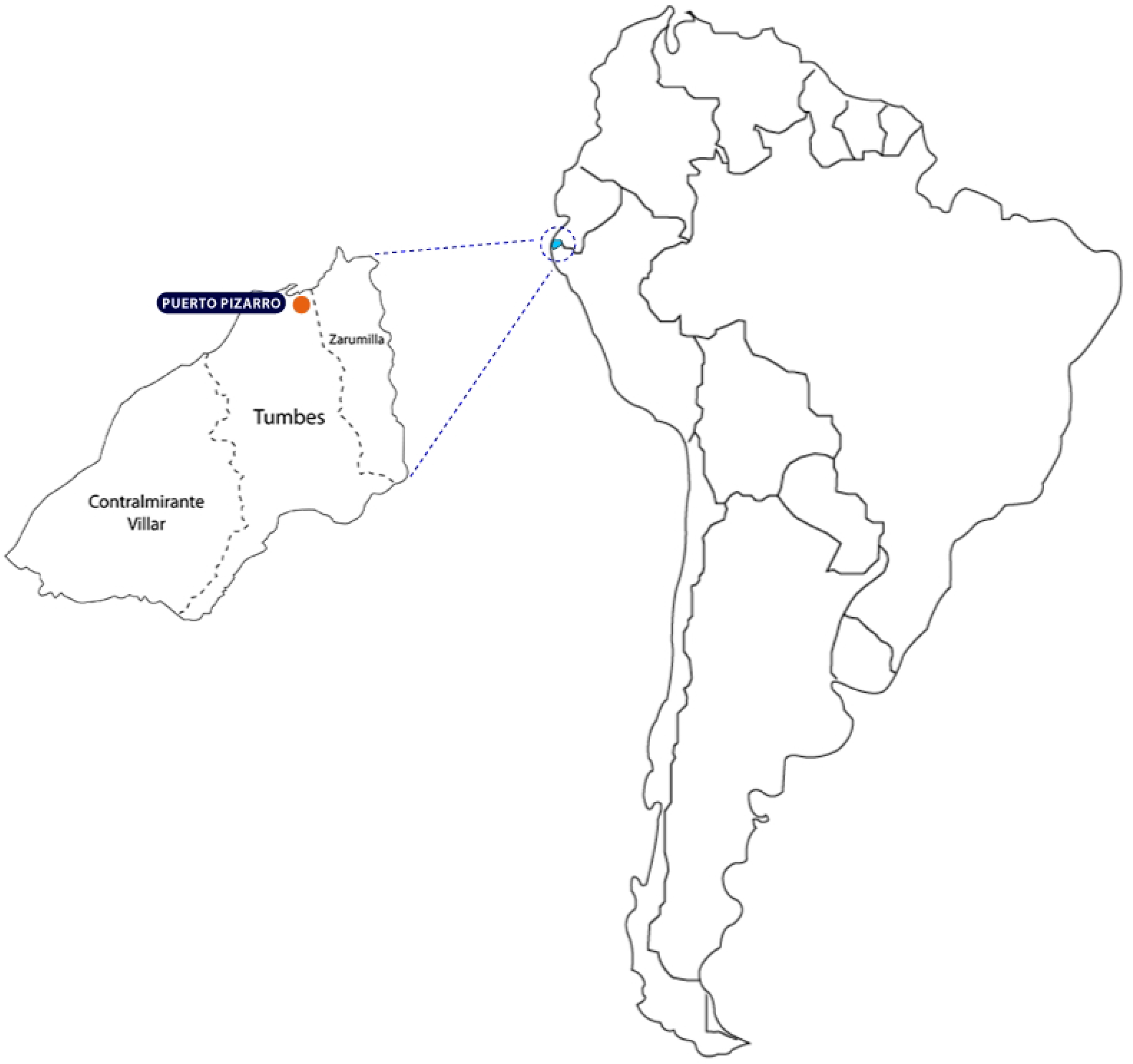
Map of Puerto Pizarro in Tumbes, Peru. “Reprinted from geogpsperu.com under a CC-BY license, with permission from Juan Pablo Suyo Pomalia, publicly available at https://www.geogpsperu.com/2019/05/america-del-sur-limites-shapefile.html.

### Phase I

A baseline census was conducted to collect data on households. After obtaining IC (13), non-medical field workers (trained by a team of epidemiologists LMMV, RGM, and PVB) performed a door-to-door survey of all individuals older than 2 years old. Individuals were evaluated to identify clinical symptoms consistent with COVID-19 disease according to the WHO (14,15). Also, sociodemographic questions previously validated by the Center for Global Health in previous epidemiological interventions were made (13,16). To ensure comprehension, illiterate individuals were included by reading aloud the IC and survey. Comorbidities defined as the presence of at least one of the following medical conditions were also identified: hypertension, diabetes, hepatic disease, renal disease, lung disease, asthma, obesity, neurological conditions, HIV, cancer, or tuberculosis (17).

### Study design and sampling

Observational, cross-sectional study. Every person in one out of every four Puerto Pizarro homes (n = 742) was asked to take part in a systematic random sampling.

### Procedures

The interviews took place between November 11th and November 30th, 2020. After enrolling all participants in each selected house, the study team took a blood sample and explained the rapid test findings to the participants. Individuals with respiratory symptoms were examined by a clinician from the study team and were registered in the Notiweb and SISCOVID databases for follow-up by the health system. Within the same house, an adult of legal age was selected and an additional molecular test (RT-PCR) was taken to look for active COVID-19 cases.

### Rapid COVID-19 tests

A lateral flow test (LFIA, lateral flow immunoassay) was used to detect the antibody positivity against SARS-CoV-2. STANDARD Q COVID-19 IgM/IgG (SD Biosensor, Republic of Korea) is a rapid chromatographic immunoassay for the qualitative detection of anti-SARS-CoV-2 nucleocapsid (N) protein antibodies and anti-SARS-CoV-2 spike RBD antibodies in human serum, plasma, or whole blood (18). The manufacturer reports 99.03 percent sensitivity (102/103) and 98.65 percent specificity (219/222). Two trained reviewers (RG and VC) read the results, and a third reviewer resolved discrepancies (VH).

### Statistical analysis

Stata v 17.0 was used to perform the statistical analysis (College Station, Texas 77845, USA). We estimated the seroprevalence ratios of chosen variables using descriptive statistics and binomial family generalized lineal models with a logarithmic link function, while controlling for other variables such as the participant’s age and gender. To account for household clustering, robust sandwich-type standard errors were utilized. Each variable was evaluated for inclusion in the final model using the log likelihood ratio. We report confidence intervals of 95% and set the level of statistical significance at p<0.05. Seroprevalence was adjusted for the test’s reported sensitivity (99.03%) and specificity (98.65%).

## Results

A total of 1916/4000 individuals older than 2 years were invited to participate, 1391/1916 (72.5 %) signed an IC. Female participants were 53.3 % (742/1,391). The participants’ mean age was 29±19 years. The majority of people lived in single families 1322/1391 (95.9 %), and had a median of four rooms and four inhabitants per home. A large proportion of participants 1287/1391 (92.5 %) had access to public potable water for a few hours (01-02 hours per day) and 1109/1391 (79.7 %) had access to hygienic services (bath).

### Seroprevalence

Anti-SARS-CoV-2 antibodies were detected in 356/1391 (25.5 %) participants: 264 IgG reactive, 78 IgM/IgG reactive, and 14 IgM reactive. After adjusting for sensitivity (99.03%) and specificity (98.65%), the calculated adjusted seroprevalence was 24,72%.

### Sociodemographic characteristics

Women had a slightly higher adjusted seroprevalence compared to men (213/356 vs 143/356 [28.01 % vs 21.18 %], p=0.005). No other demographic characteristics were associated with SARS-CoV-2 rapid test seropositivity (Table 1).

**Table 1.**
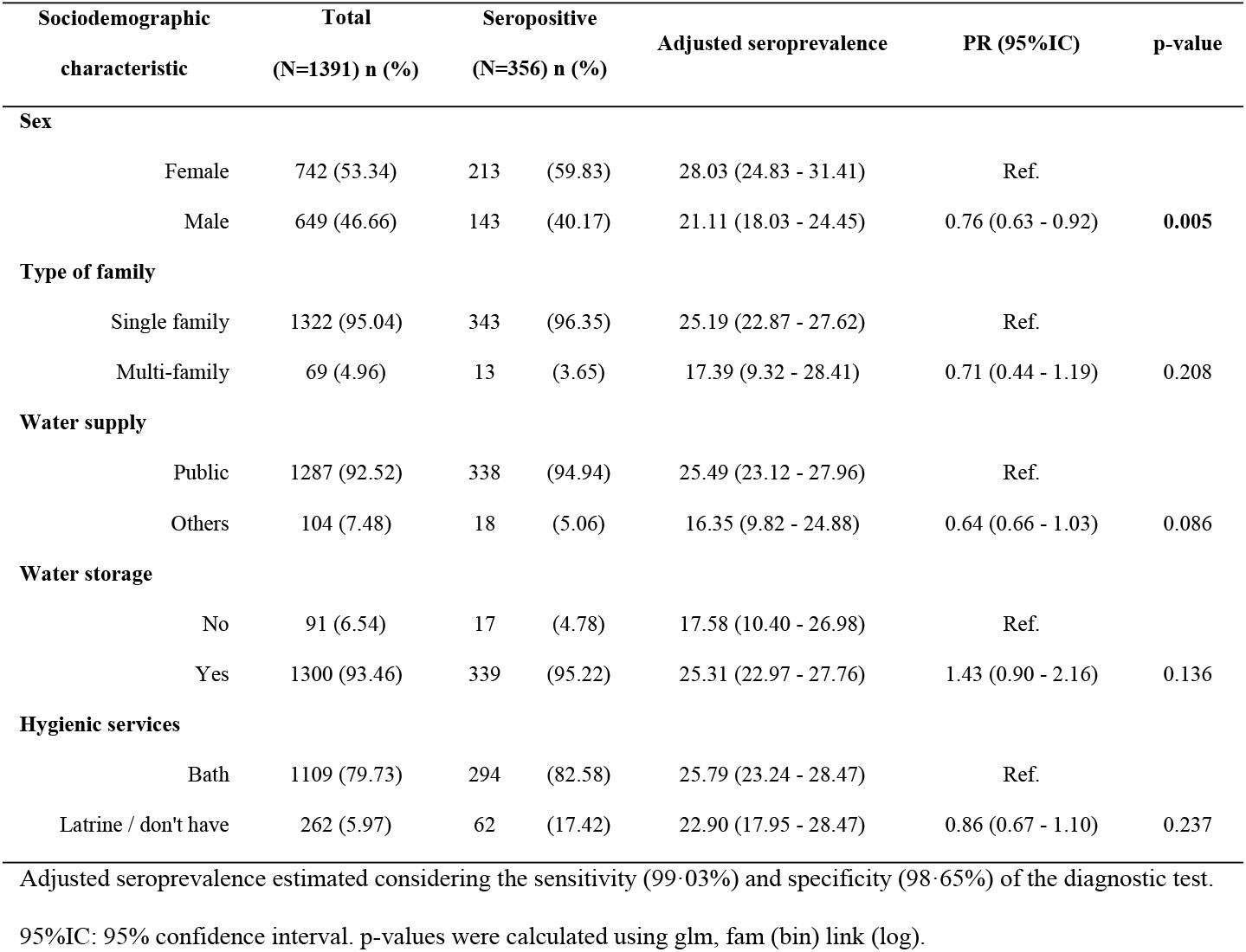
Seroprevalence adjusted by participant characteristics and associated factors.

As illustrated in Fig 2, the age groups with the highest prevalence of IgG seropositive individuals were those under 16 years old, 41-50 years old, and 66-75 years old.

**Fig 2.**
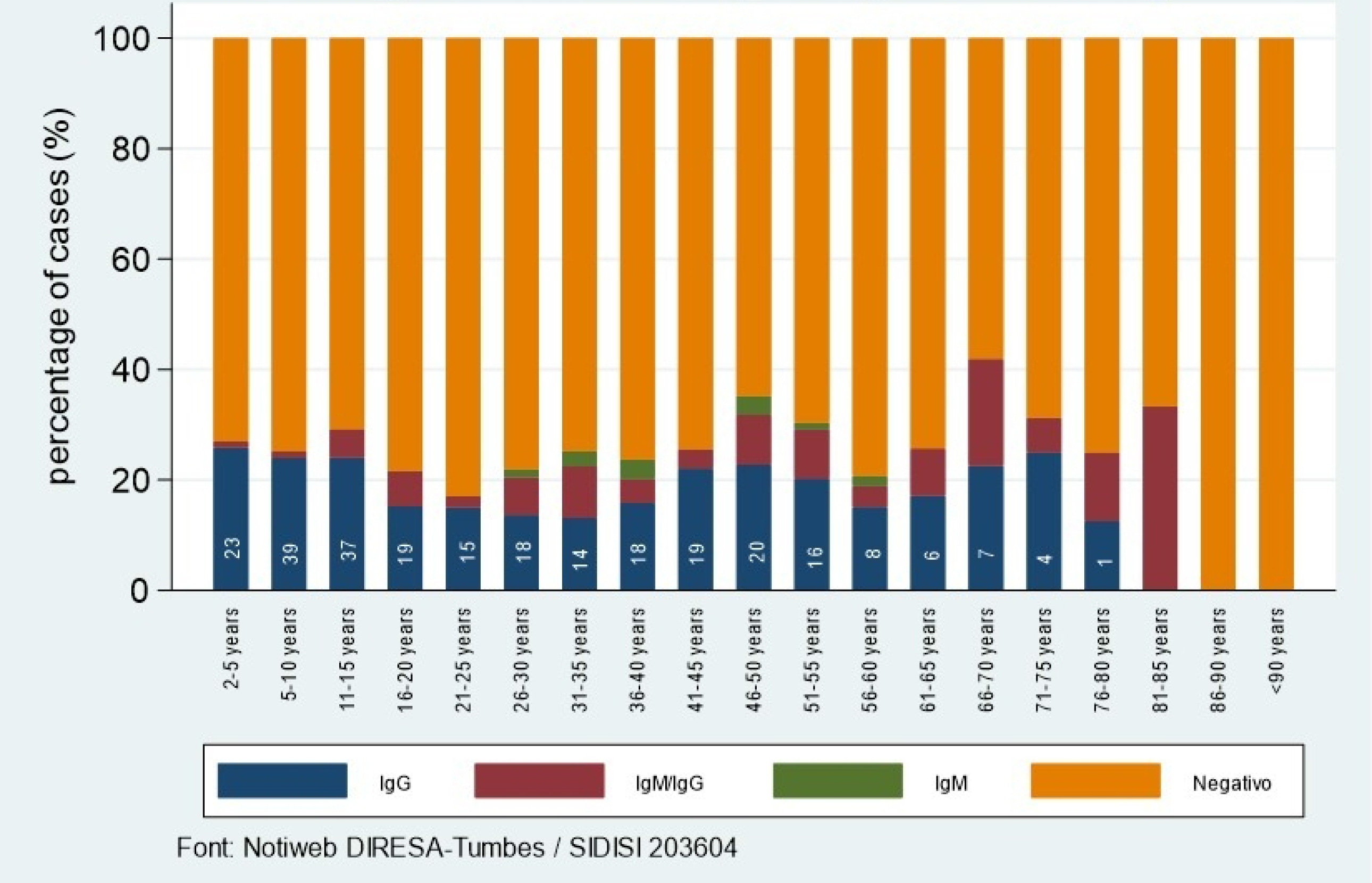
Seronegative and seropositivity in Puerto Pizarro. Cases in the participants of study

The RT-PCR test revealed no active cases and no deaths were reported among the participants. On the other hand, 66.3% of IgM and IgM/IgG positive patients were asymptomatic (Table 2).

**Table 2.** Number of symptomatic and asymptomatic cases in the seropositive population.

| Age group | IgM positive |  | IgM/IgG positive |  | IgG positive |  | All seropositives |  | Total |
| --- | --- | --- | --- | --- | --- | --- | --- | --- | --- |
|  | No | Yes | No | Yes | No | Yes | No | Yes |  |
| 2-5 years | 0 | 0 | 1 | 0 | 20 | 3 | 21 | 3 | 24 |
| 6-10 years | 0 | 0 | 2 | 0 | 34 | 5 | 36 | 5 | 41 |
| 11-15 years | 0 | 0 | 7 | 1 | 29 | 8 | 36 | 9 | 45 |
| 16-20 years | 0 | 0 | 4 | 4 | 14 | 5 | 18 | 9 | 27 |
| 21-25 years | 0 | 0 | 1 | 1 | 11 | 4 | 12 | 5 | 17 |
| 26-30 years | 2 | 0 | 5 | 4 | 10 | 8 | 17 | 12 | 29 |
| 31-35 years | 3 | 0 | 4 | 6 | 11 | 3 | 18 | 9 | 27 |
| 36-40 years | 3 | 1 | 3 | 2 | 15 | 3 | 21 | 6 | 27 |
| 41-45 years | 0 | 0 | 3 | 0 | 10 | 9 | 13 | 9 | 22 |
| 46-50 years | 1 | 2 | 7 | 1 | 16 | 4 | 24 | 7 | 31 |
| 51-55 years | 0 | 1 | 5 | 2 | 10 | 6 | 15 | 9 | 24 |
| 56-60 years | 0 | 1 | 2 | 0 | 5 | 3 | 7 | 4 | 11 |
| 61-65 years | 0 | 0 | 2 | 1 | 4 | 2 | 6 | 3 | 9 |
| 66-70 years | 0 | 0 | 5 | 1 | 5 | 2 | 10 | 3 | 13 |
| 71-75 years | 0 | 0 | 0 | 1 | 2 | 2 | 2 | 3 | 5 |
| 76-80 years | 0 | 0 | 0 | 1 | 0 | 1 | 0 | 2 | 2 |
| 81-85 years | 0 | 0 | 1 | 1 | 0 | 0 | 1 | 1 | 2 |
| 86-90 years | 0 | 0 | 0 | 0 | 0 | 0 | 0 | 0 | 0 |
| <90 years | 0 | 0 | 0 | 0 | 0 | 0 | 0 | 0 | 0 |
|  | 9 | 5 | 52 | 26 | 196 | 68 | 257 | 99 | 356 |

In a bivariate analysis, numerous symptoms such as fever (PR: 1.89 95% IC[1.44 - 2.48]), general discomfort (PR: 1.67, 95% IC [1.23 - 2.26], 0.001), cough (PR:2.00; 95% IC [1.60 - 2.50]), nasal congestion (PR: 1.46, 95% IC [1.03 - 2.09]), respiratory distress (PR: 1.64, 95% IC [1.04 - 2.56]), anosmia (PR: 1.78, 95% IC [1.01 - 3.14]) and ageusia (PR: 2.31, 95% IC [1.48 – 3.61]) were associated with SARS-CoV-2 seropositivity. Cough was the unique symptom associated with seropositivity in a multiple analysis (PR: 1.78 95 percent IC [1.24 - 2.57]) (Table 3). There were no comorbidities associated with seropositivity.

**Table 3.** Self-reported symptoms between seropositive and seronegative participants.

| Covid-19 symptoms | Positives |  | Negatives |  | PR (CI 95%) | p-value* | PRA (CI 95%) | p-value** |
| --- | --- | --- | --- | --- | --- | --- | --- | --- |
|  | n=356 |  | n=1035 |  |  |  |  |  |
|  | yes | no | yes | No |  |  |  |  |
| Fever | 34 | 322 | 42 | 993 | 1.89 (1.44 - 2.48) | <0.001 | 1.29 (0.84 - 1.98) | 0.254 |
| General Discomfort | 28 | 328 | 42 | 993 | 1.67 (1.23 - 2.26) | 0.001 | - | - |
| Cough | 53 | 303 | 63 | 972 | 2.00 (1.60 - 2.50) | <0.001 | 1.78 (1.24 - 2.57) | 0.002 |
| Sore Throat | 39 | 317 | 126 | 909 | 0.95 (0.71 - 1.27) | 0.716 | - | - |
| nasal congestion | 21 | 335 | 38 | 997 | 1.46 (1.03 - 2.09) | 0.036 | 1.08 (0.67 - 1.75) | 0.742 |
| Respiratory Distress | 18 | 338 | 12 | 1023 | 1.64 (1.04 - 2.56) | 0.031 | 0.99 (0.53 - 1.88) | 0.999 |
| Diarrhea | 3 | 353 | 12 | 1023 | 0.81 (0.29 - 2.23) | 0.676 | - | - |
| Sickness | 3 | 353 | 4 | 1031 | 1.73 (0.73 - 4.10) | 0.209 | - | - |
| Headache | 22 | 334 | 37 | 998 | 1.54 (1.09 - 2.17) | 0.014 | - | - |
| Irritability | 2 | 354 | 5 | 1030 | 1.15 (0.35 - 3.74) | 0.811 | - | - |
| Muscle pain | 4 | 352 | 6 | 1029 | 1.62 (0.75 - 3.48) | 0.215 | - | - |
| Abdominal pain | 1 | 355 | 2 | 1033 | 1.34 (0.27 - 6.69) | 0.716 | - | - |
| Chest pain | 4 | 352 | 0 | 1035 | - | - | - | - |
| Join pain | 2 | 354 | 0 | 1035 | - | - | - | - |
| Anosmia | 7 | 349 | 9 | 1026 | 1.78 (1.01 - 3.14) | 0.046 | - | - |
| Ageusia | 9 | 347 | 7 | 1028 | 2.31 (1.48 - 3.61) | <0.001 | 1.57 (0.78 - 3.15) | 0.202 |

\* p-value were calculated with glm, fam(bin) link(log)
\*\* p-value were calculated glm, fam(bin) link(log), adjusted by anosmia, fever, cough, respiratory distress and nasal congestion

## Discussion

In Tumbes, the first wave of COVID-19 took place between March 16, 2020 and the first days of December, 2020. The present cross-sectional serological survey took place eight months after the first case of COVID-19 was detected in Tumbes, between November 11 and November 30. Serology was performed during the epidemiological weeks (EW) 45–48, posterior to the first official peak of cases and mortality in Tumbes. Therefore, this study will help to understand how the first wave of COVID-19 affected this population’s serology. Tumbes had an adjusted seroprevalence of 24.72 %, whereas the seroprevalence in other regions of Peru’s interior ranged between 10.2 and 70 % (7–9).

Seropositivity in the present study was slightly higher compared to similar studies conducted in other developing settings such as Nairobi (22.7% [95% CI: 18.0-27.7]) or Palestine (24.0% [95% CI: 21.7-26.5]) (19,20). However, the high seroprevalence could be associated with the behaviors of people regardless of whether they belong to a developed or developing country. After the first 8 months of the pandemic, the Netherlands had 19.5 % SARS-CoV-2 seroprevalence. The factors associated with exposure to the virus were no social distancing and a high number and duration of contacts (21). An epidemiological study in Lima, Peru found that a gradual increase in the overcrowding index was associated with a higher seroprevalence of COVID-19 (aPR = 199 [95 percent CI 141-281, p<0.001] in the fourth quartile). However, the number of rooms and family members were not associated with positive seroprevalence in our study. The informal economy, on the other hand, required people to leave home every day to work, exposing them to COVID-19 during the first wave, which is consistent with what was observed in Lima, Peru, where a gradual decrease in socioeconomic status was associated with a higher seroprevalence (aPR = 3.41 [95 percent CI: 1.90-6.12, p<0.001] in low socioeconomic status) (9). It is known that the scarcity of potable water (available 1 or 2 hours per day), the use of tanker trucks, commercial and unlicensed suppliers, informal arrangements with neighbors (22,23) and restricted sanitary practices, such as hand washing, significantly reduce the effectiveness of COVID-19 prevention and control measures. But our study found no association between positive seroprevalence and water supply, water resources, or sanitation services (7,8,14).

When adjusted by participant characteristics, women had higher adjusted seroprevalence compared to men (213/356 vs 143/356 [28.01 % vs 21.18 %], p=0.005). Similar studies had reported no association between sex and seroprevalence during the first wave of COVID-19 (24–26). This link is plausible due to mothers’ exposure to the virus while market shopping. In fact, according to an epidemiological report done by the Peruvian Ministry of Health, up to 86% of merchants in Peruvian markets had COVID-19 as of June 2, 2020 (27).

In order to restrict the spread of SARS-CoV-2, it is crucial to determine which age group is continually exposed to the virus. In our based-population study, more than 20% of IgG seropositive cases belong to the age groups under 16 years old, followed by 41-55 years old, and 66-75 years old (Fig 2), assuming that many asymptomatic individuals were not detected when they acquired the virus. In Iquitos (Jungle, Peru) study findings, a higher seroprevalence was observed in the age group younger than 12 years old (26), in Saudi Arabia (ASIA), the seroprevalence of SARS-CoV-2 was higher in the 1 to 18-year-old group (24); in both studies most of the seropositive children were asymptomatic. Children, despite being asymptomatic, are just as susceptible to COVID-19 as adults. One possible explanation is that SARS-CoV-2 uses ACE2 as a viral receptor, whose tissue distribution and binding capacity may be lower in children compared to adults (28). The evidence reveals that the viral load does not differ considerably between symptomatic and asymptomatic individuals; therefore, asymptomatic individuals represent a source of the transmission of the virus. (29,30).

Fever, sore throat, and cough were the most frequently reported symptoms in COVID-19 in earlier research in Lima-Peru, occurring in 40% to 50% of seropositive symptomatic patients (31). Dysosmia (PR = 1.69), chest pain (PR = 1.49), back pain (PR = 1.45), cough (PR = 1.44), fever (PR = 1.41), and general malaise (PR = 1.27) were also shown to be linked with an increased prevalence of SARS-CoV-2 seropositivity in Lambayeque’s study (8). Our findings corroborated those seen in national and international reports (32,33); cough was the single symptom associated with a positive COVID-19 quick test, suggesting that the disease’s behavior may differ between different places.

Asymptomatic individuals (34,35) with recent infection were 66.3% (IgM and IgM/IgG) across all age groups in our study (see Table 2). This exceeds previously reported evidence suggesting that asymptomatic transmission of SARS-CoV-2 may account for between 30–59% of all infections. (36). Also, infected people can be asymptomatic carriers, complicating their identification by health systems and being the principal viral transmitter inside their homes. (37)(38). In this context, community screening is required to determine the true dynamics of infection transmission and to adjust control measures through population-based tactics such as active search for suspected cases (door-to-door registration) and early case confirmation with antigen rapid testing.

Our study had a limitation. We did not perform RT-PCR molecular tests on seronegative patients, and hence, we were unable to estimate the infection rate among those who were asymptomatic and seronegative participants.

Extrapolating the adjusted prevalence from our study (24.72 per 1000 people) to the region of Tumbes (N = 251,541), about 62,180 individuals were infected during the first wave, too high for optimal primary care in health facilities. The number of COVID-19 cases in the region exceeded what has been reported in regional health bulletins (39). Multiple variables may have contributed to the underreporting: the presence of primarily symptomatic patients at health care facilities; inadequate use of social networks; scarce eHealth tools in government facilities; unused emergency telephone numbers; and the fear of dying if referred to the hospital. In future studies, home-made antigenic rapid tests for early viruses using smartphone technology could be a plausible solution to be explored.

Some key behaviors related with the risk of COVID-19 infection, such as rigorous home isolation and recent close contact with positive individuals, were not analyzed and may be investigated in future research in Tumbes. Diaz-Velez et al, in Lambayeque’
ss study, demonstrated that individuals who complied with rigorous home isolation had a 20% lower likelihood of testing positive for SARS-CoV-2 (PR = 0.80). The odds of testing positive went up by 60% (PR = 1.60) in contact with patients with acute respiratory illness, 51% (PR = 1.51) in contact with a confirmed case in the past 14 days, and 26% (PR = 1.26) in visits to the market (8).

## Conclusions

This study shows a high prevalence of SARS-CoV-2 antibodies after the first wave of COVID-19 in Tumbes, which can provide an estimation of the infection attack rate during this period. A multidisciplinary effort must be made to understand the development of the disease and the behavior of the communities around COVID-19. So far, there is an urgent need to understand the real situation of COVID-19 in vulnerable populations to improve surveillance and control programs.

## Data Availability

All relevant data are within the manuscript and its Supporting Information files.

## Acknowledgments

We thank the villagers of Puerto Pizarro. LMMV and team are grateful to all the regional authorities from the Regional Government of Tumbes; from the Local Government and the Regional Directorate of Health-Tumbes V Jimenez, and J Arias. We are grateful to the members of the Executive Office of Epidemiology N Julca, and L Arevalo. LM expresses gratitude to the members of the Neuroepidemiology and Science of Life group from Universidad Cesar Vallejo. LM thanks the Universidad Nacional de Tumbes and the 1era Brigada de Infantería del Ejército Peruano for their assistance with field staff mobilization and safety. Jenny Chirinos and Angie Toledo are doctoral students in the Epidemiological Research Doctorate at Universidad Peruana Cayetano Heredia under FONDECYT/CIENCIACTIVA scholarship EF033-235-2015 and supported by training grant D43 TW007393 awarded by the Fogarty International Center of the US National Institutes of Health.

## Footnotes

The statements contained in this document are those of the authors and should not be construed as official points of view of the Universidad Cesar Vallejo, Universidad Nacional de Tumbes, or other organizations mentioned.

